# Troponin T and Neurofilament Light Chain Levels as Complementary Biomarkers of Disease Accumulation and Aggressiveness in Amyotrophic Lateral Sclerosis

**DOI:** 10.64898/2026.05.17.26353398

**Authors:** Julia Meyer, Sofia Waldorf, Janina von der Gablentz, Torsten Grehl, Huelya Nazlican, Thomas Meyer, Julian Grosskreutz, Patrick Weydt, Sarah Bernsen

**Affiliations:** University of Luebeck, Clinic of Neurology, Precision Neurology of Neuromuscular and Motor Neuron Disease, Luebeck, Germany; Clinic for Parkinson’s Disease, Sleep Disorders, and Movement Disorders, University Hospital Bonn, Bonn, Germany; Department of Neurology, Alfried Krupp Hospital, Essen, Germany; Department of Neurology, Center for ALS and Other Motor Neuron Disorders, Charite - Universitaetsmedizin Berlin, Corporate Member of Freie Universitaet Berlin, Humboldt-Universitaet zu Berlin and Berlin Institute of Health, Berlin, Germany; Ambulanzpartner Soziotechnologie APST GmbH, Berlin, Germany; University of Luebeck, Cluster for Precision Medicine in Inflammation, Luebeck, Germany

## Abstract

**Objectives:** Amyotrophic lateral sclerosis (ALS) is a clinically heterogeneous neurodegenerative disease requiring reliable biomarkers to improve patient stratification and trial design. While serum neurofilament light chain (sNfL) reflects neuroaxonal stress and disease aggressiveness, troponin T (TnT) may capture complementary aspects of neuromuscular involvement. We assessed the associations of TnT and sNfL with D50-derived measures of disease aggressiveness (D50) and disease accumulation (rD50) in ALS.

**Material and Methods:** In this retrospective observation, TnT and sNfL levels from ALS patients in two independent German cohorts were analyzed using the D50 disease progression model; discovery cohort (Essen, n =433) and validation cohort (Bonn, n =185).

**Results:** In both cohorts TnT demonstrated a robust correlation with rD50-defined phases across all aggressiveness subgroups (p<0.001). There was no consistent pattern regarding sNfL and the rD50 phases.

sNfL concentrations demonstrated a significant and inverse correlation with D50 applied for all disease aggressiveness subgroups (p<0.001). Correlations of TnT levels with D50 disease aggressiveness groups were generally less strong and inconsistent between the two cohorts. In the discovery cohort only low aggressiveness subgroups correlated significantly (p<0.001), intermediate aggressiveness subgroups showed only a weak correlation (p<0.05) with TnT levels. High disease aggressiveness subgroups showed no significant correlation with TnT.

**Conclusion:** In application of the D50 disease progression model, TnT was strongly associated with disease accumulation (rD50) across all disease phases, independent of disease aggressiveness (D50), whereas sNfL robustly reflected disease aggressiveness but not overall disease burden.

These complementary biomarker profiles highlight the value of an integrated approach for refined disease stratification in ALS. Combining TnT and sNfL may enhance clinical decision-making, improve monitoring of disease progression and treatment response, and support optimized clinical trial design.

## 1. Introduction

Amyotrophic lateral sclerosis (ALS) is a fatal neurodegenerative disease marked by progressive degeneration of upper and lower motor neurons, leading to muscle weakness, atrophy, and spasticity^1^. Despite ongoing research, effective treatments remain scarce. The clinical trajectory of ALS is highly heterogeneous, with wide interindividual variability in age of onset, rate of progression, symptom distribution, and survival^2^. Understanding and predicting the individual course of the disease is a central challenge in ALS management. Therefore, reliable surrogate markers are urgently needed to enable optimized stratification for future treatments and clinical trial designs^3^.

Functional decline is conventionally assessed using the revised ALS Functional Rating Scale (ALSFRS-R)^4^. The ALSFRS-R is inherently susceptible to measurement variability that does not necessarily reflect true biological disease progression. Given its ordinal and partly subjective structure, small changes in score may arise from intra- and inter-rater differences or transient patient-related factors, thereby limiting its sensitivity and reliability as a sole indicator of disease dynamics^5 6^.

This presents a considerable challenge for the design of clinical studies and is increasingly taken into account in both stratification and analytical approaches.

The disease progression rate (DPR), derived from the monthly decline in the ALSFRS-R, is widely used as a quantitative indicator of clinical disease aggressiveness in ALS; however, it has important methodological limitations. Specifically, the DPR assumes a linear decline in ALSFRS-R sum scores, whereas large longitudinal studies suggest that functional decline follows a rather curvilinear trajectory over the individual disease course^7^. Moreover, calculating progression based on a single ALSFRS-R assessment renders the measure susceptible to intra- and inter-rater variability, potentially affecting its reliability ^8 9^.

The D50 model of ALS was developed to pragmatically mitigate the limitations of conventional clinical measures and account for different progression types. The model provides a framework for interpreting biomarker signals that is distinct from the accumulation of disease burden^10 11^. The model uses an individual sigmoidal fitting from health to function loss, based on the longitudinally collected ALSFRS-R for each patient. It allows the description of the aggressiveness of the disease and the stage of individual progression.

Valid biomarkers provide important insights into the pathophysiology of the disease, but they also serve as markers of disease phase, progression, and therapeutic response.

Neurofilament elevations in serum and cerebrospinal fluid (CSF) are markers of neuroaxonal stress, increase early in the disease course of ALS with higher concentrations in CSF than in serum and remain stable over time. Neurofilaments primarily reflect upper motor neuron pathology and serve as proxies of disease aggressiveness and survival^11^. To date, neurofilaments represent the most important fluid biomarker in ALS for both disease progression and treatment response^12 13^.

Two species of measurable neurofilaments have been established as biomarkers: neurofilament light chain (NfL) and phosphorylated neurofilament heavy chain (pNfH). NfL can be reliably quantified in both CSF and serum, reflecting its ability to cross the blood–brain barrier and its suitability for minimally invasive assessment^14^. In contrast, pNfH is most commonly measured in CSF.

Application of the D50 model to both pNfH and NfL in CSF demonstrates the potential of neurofilaments to capture disease aggressiveness independent of disease accumulation while remaining stable throughout the disease course ^15 16^. Serum NfL (sNfL) is prone to more fluctuations compared to CSF-NfL; however, overall sNfL levels remain relatively stable throughout the disease course ^10 17^.

Troponin T (TnT), a well-established cardiac biomarker, is being repurposed in neuromuscular disorders, including ALS. Elevated TnT levels are common in ALS and likely reflect regenerative and/or degenerative skeletal muscle activity and improve diagnostic accuracy for ALS in combination with sNFL ^18^-^20^.

In untreated patients, TnT increases over time, correlating with functional decline and suggesting its use as a marker of disease accumulation ^21^. The stabilization of TnT levels under Tofersen treatment further supports its potential utility as a treatment-response biomarker ^22^. While its prognostic value remains under investigation, the accessibility of TnT and its complementary nature to NfL make it a promising candidate for disease monitoring.

The D50 model enables independent quantification of disease aggressiveness and disease accumulation, which is essential to understand the validity of a biomarker, particularly relevant for clinical trials where patients progress through different phases.

NfL has emerged as a robust aggressiveness-related marker within this model but not for cumulative functional loss. Against this background, we applied the D50 framework to determine whether TnT also corresponds to disease aggressiveness or rather compliments NfL and might capture a distinct aspect of disease biology indicative of disease accumulation.

## 2. Methods

### 2.1 Study design

We performed a retrospective analysis of two patient cohorts from specialized ALS centers of the Alfried Krupp Hospital Essen (discovery cohort) and the University Hospital Bonn (validation cohort). All patients had a confirmed ALS diagnosis and at least two clinical visits to allow stringent D50 modeling. Patients in the discovery cohort were seen in the ALS clinic between January 10, 2023, and April 1, 2025, the validation cohort had visits between January 1, 2020, and December 1, 2023. Data were extracted in both cohorts from routine clinical records and included demographic information (age, sex), clinical variables (ALSFRS-R, site and date of onset), and biomarker levels (TnT, sNfL) taken at each visit.

### 2.2 Laboratory Markers

sNfL concentrations (Essen) were measured at the ALS Center Berlin by means of the SIMOA, using the NfL Advantage Kit (Quanterix Inc., USA). They were prospectively obtained via the APST registry and its “NfL-ALS” substudy. The study protocol for the APST registry and the substudy “NfL-ALS” were approved by the Medical Ethics Committee of Charité-Universitätsmedizin Berlin, Germany, under numbers EA2/168/20 and EA1/219/15.

sNfL levels (Bonn) were measured at the University Medical Center Ulm using a standardized ELISA protocol^23^.

High-sensitivity assays for TnT were performed using Electrochemiluminescence Immunoassay (ECLIA) at the central laboratory at Alfried Krupp Hospital (discovery cohort) and in a fully accredited commercial laboratory (Labor Volkmann, Karlsruhe) (validation cohort).

No additional informed consent or ethical approval was required from the institutional ethics review board in Bonn, for the underlying data of this analysis was fully pseudonymized routine clinical data (Ethics board approval letter 2025/069W, Bonn).

### 2.3 D50 Model

The sigmoidal fit of the D50 model can be described with two parameters, first, D50 as time in months since initial symptom onset to the time point of 50% motor function loss, as loss of 24 points in the 48-point ALSFRS-R score and second, dx as time constant of functional decline, as steepness of the curve. As described before, D50 and dx correlate directly in different cohorts^24 25 26^.

The use of the D50 model enables robust comparison across heterogeneous patient cohorts by quantifying disease aggressiveness independently of disease duration at the time of assessment. Unlike conventional cross-sectional metrics, this approach accounts for individual differences in baseline status and temporal disease evolution, allowing patients at different clinical stages to be aligned on a common progression scale. Consequently, stratification the patients participating in this analysis into disease aggressivenss subgroups of high (D50 < 20 months), intermediate (20 ≤ D50 < 40 months), and low (D50 ≥ 40 months) facilitates meaningful comparisons between cohorts by reducing bias introduced by variability in disease onset and observation time.

By normalizing the real-time D50 value the relative D50 (rD50) can be calculated, a dimensionless parameter that maps disease progression on an open-ended scale. An rD50 value of 0 is the time of disease onset, while a value of 0.5 represents the time point of 50% motor function loss, that is the D50.

This parameter can be calculated for any given time point during the disease course, enabling time-normalized staging independent of individual progression speed.

With the help of rD50, we can categorize three disease phases: Phase I is designated as the early semi-stable phase and is characterized by rD50 values ranging from 0 to 0.25. Phase II, the early progressive stable phase, is characterized by rD50 values ranging from 0.25 to 0.5. Phase III/IV, the late progressive and late stable phase, is defined by rD50 values ≥0.5.

The D50 model allows the calculation of two further descriptors of local disease activity at any given time point. The calculated functional loss rate (cFL), expressed as points lost per month and the steepness of the sigmoidal curve, and the calculated functional state (cFS), representing the estimated ALSFRS-R score at a specific time point on the individual disease course.

As shown by Meyer et al. 2025 the D50 parameter may be superior to cFL and DPR for quantifying disease aggressiveness, therefore our analysis focused primarily on D50.

### 2.4 Statistics

The statistical analysis and graphical visualization of the datasets were conducted using RStudio open-source software (Version 2023.06.01, Posit Software, PBC, Boston, MA, United States) and the R programming language on a macOS Sequoia 15.5. The assessment of normality was conducted through the implementation of the log-10 transformation and the Shapiro-Wilk test, thereby enabling the implementation of parametric testing for the following variables: D50, sNfL levels and TnT levels.

Linear regression models were applied for the two biomarker variables, ln(NfL) and ln(TnT), together with ln(D50) and ln(rD50) as the dependent variable.

Differences in sNfL and TnT concentrations within each ALS cohort were assessed using one-way analysis of covariance (ANCOVA), adjusting for age at onset, sex, site of onset, disease aggressiveness, and disease accumulation, followed by post hoc Wilcoxon rank-sum tests.

## 3. Results

### 3.1. Patient characteristics

Demographic, clinical data and biomarker levels of the two cohorts are shown in Table 1 (Essen; discovery cohort) and Table 2 (Bonn; validation cohort) stratified by D50-derived subgroups according to disease accumulation (rD50) and disease aggressiveness (D50).

**Table 1:**
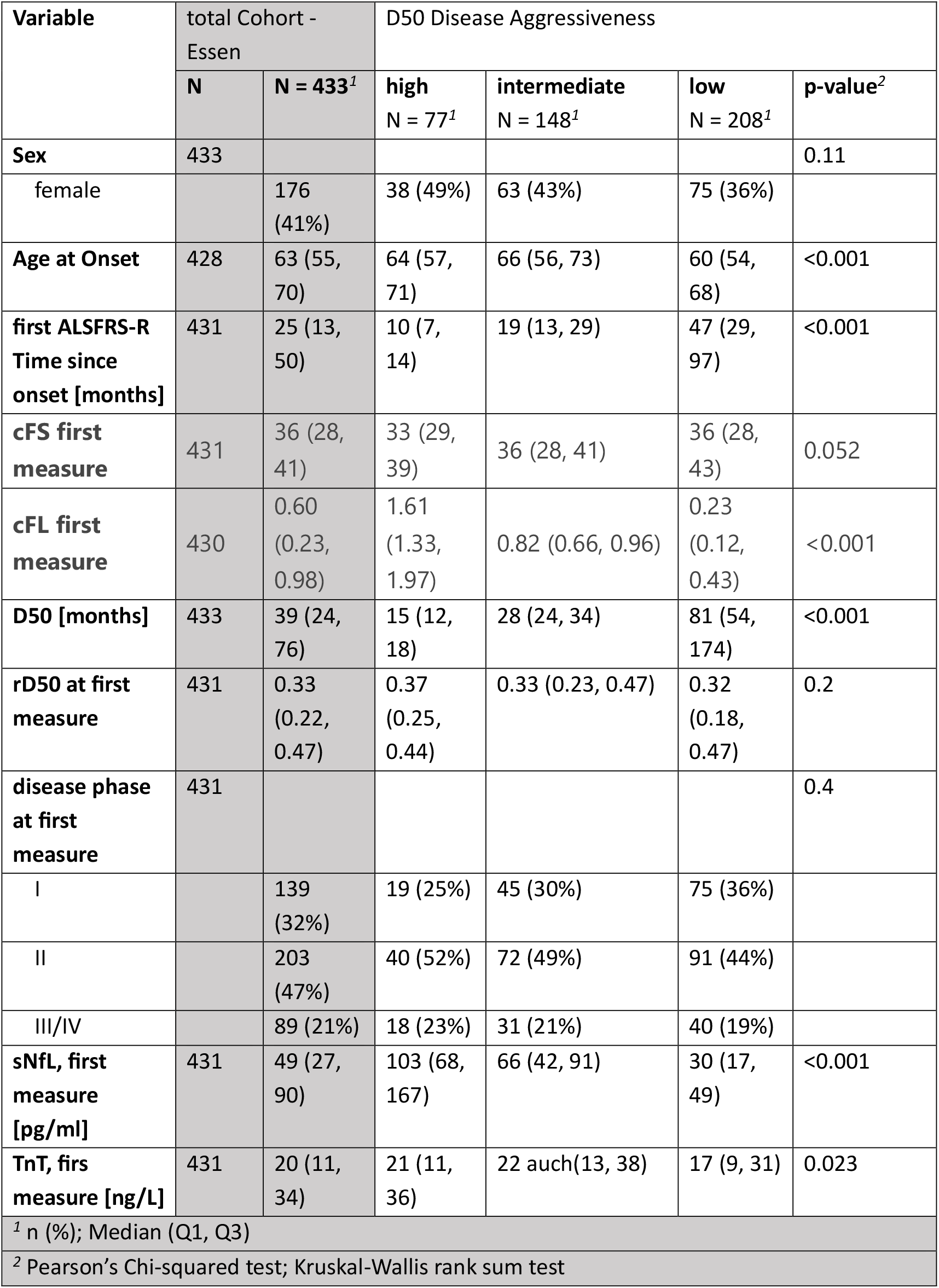
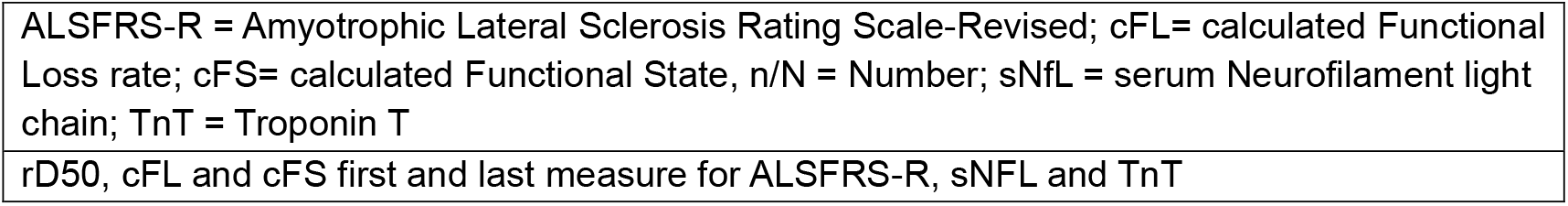
Demographic and clinical characteristics of the discovery cohort (Essen, n = 433)

| Variable | total Cohort - Essen |  | D50 Disease Aggressiveness |  |  |  |
| --- | --- | --- | --- | --- | --- | --- |
|  | N | N = 433<sup>1</sup> | high<br>N = 77<sup>1</sup> | intermediate<br>N = 148<sup>1</sup> | low<br>N = 208<sup>1</sup> | p-value<sup>2</sup> |
| **Sex** | 433 |  |  |  |  | 0.11 |
| female |  | 176 (41%) | 38 (49%) | 63 (43%) | 75 (36%) |  |
| **Age at Onset** | 428 | 63 (55, 70) | 64 (57, 71) | 66 (56, 73) | 60 (54, 68) | <0.001 |
| **first ALSFRS-R Time since onset [months]** | 431 | 25 (13, 50) | 10 (7, 14) | 19 (13, 29) | 47 (29, 97) | <0.001 |
| **cFS first measure** | 431 | 36 (28, 41) | 33 (29, 39) | 36 (28, 41) | 36 (28, 43) | 0.052 |
| **cFL first measure** | 430 | 0.60 (0.23, 0.98) | 1.61 (1.33, 1.97) | 0.82 (0.66, 0.96) | 0.23 (0.12, 0.43) | <0.001 |
| **D50 [months]** | 433 | 39 (24, 76) | 15 (12, 18) | 28 (24, 34) | 81 (54, 174) | <0.001 |
| **rD50 at first measure** | 431 | 0.33 (0.22, 0.47) | 0.37 (0.25, 0.44) | 0.33 (0.23, 0.47) | 0.32 (0.18, 0.47) | 0.2 |
| **disease phase at first measure** | 431 |  |  |  |  | 0.4 |
| I |  | 139 (32%) | 19 (25%) | 45 (30%) | 75 (36%) |  |
| II |  | 203 (47%) | 40 (52%) | 72 (49%) | 91 (44%) |  |
| III/IV |  | 89 (21%) | 18 (23%) | 31 (21%) | 40 (19%) |  |
| **sNfL, first measure [pg/ml]** | 431 | 49 (27, 90) | 103 (68, 167) | 66 (42, 91) | 30 (17, 49) | <0.001 |
| **TnT, first measure [ng/L]** | 431 | 20 (11, 34) | 21 (11, 36) | 22 auch(13, 38) | 17 (9, 31) | 0.023 |

<sup>1</sup> n (%); Median (Q1, Q3)
<sup>2</sup> Pearson's Chi-squared test; Kruskal-Wallis rank sum test
rD50, cFL and cFS first and last measure for ALSFRS-R, sNFL and TnT

**Table 2:** Demographic and clinical characteristics of the validation cohort (Bonn).

rD50, cFL and cFS first and last measure for ALSFRS-R, sNFL and TnT
| Variable | total Cohort - Bonn |  | splitted into D50 Disease Aggressiveness |  |  |  |
| --- | --- | --- | --- | --- | --- | --- |
|  | N | N = 185<sup>1</sup> | high<br>N = 48<sup>1</sup> | intermediate<br>N = 63<sup>1</sup> | low<br>N = 74<sup>1</sup> | p-value<sup>2</sup> |
| **Sex** | 185 |  |  |  |  | >0.9 |
| female |  | 86 (46%) | 22 (46%) | 30 (48%) | 34 (46%) |  |
| **Age at Onset, years** | 185 | 60 (53, 69) | 64 (55, 73) | 62 (56, 70) | 58 (51, 66) | 0.018 |
| **first ALSFRSR Time since onset [months]** | 185 | 18 (10, 35) | 7 (5, 11) | 17 (11, 20) | 45 (25, 68) | <0.001 |
| **D50 [months]** | 185 | 34 (20, 62) | 14 (12, 17) | 30 (25, 35) | 64 (54, 104) | <0.001 |
| **rD50 at first ALSFRS-R** | 185 | 0.29 (0.20, 0.39) | 0.26 (0.18, 0.36) | 0.27 (0.20, 0.38) | 0.32 (0.21, 0.40) | 0.4 |
| **Disease phase first ALSFRS-R** | 183 |  |  |  |  | 0.7 |
| I |  | 70 (38%) | 20 (43%) | 25 (40%) | 25 (34%) |  |
| II |  | 96 (52%) | 23 (49%) | 34 (54%) | 39 (53%) |  |
| III/IV |  | 17 (9.3%) | 4 (8.5%) | 4 (6.3%) | 9 (12%) |  |
| **sNfL, first measure [pg/ml]** | 185 | 83 (52, 129) | 124 (88, 202) | 100 (63, 137) | 53 (32, 77) | <0.001 |
| **TnT, first measure [ng/L]** | 176 | 17 (9, 29) | 18 (10, 34) | 17 (8, 25) | 18 (9, 32) | 0.8 |

<sup>2</sup> Pearson's Chi-squared test; Kruskal-Wallis rank sum test; Fisher's exact test

Sex distribution was similar between the discovery (n=433) and validation cohort (n=185) (p = 0.21). Patients in the discovery cohort were older with a median age at onset of 63 years (interquartile range (IQR) 55–70) vs. 60 years (IQR 53-69) (p=0.052) (Table 3). For comparison purposes the traditional metrics (DPR and ALSFRS-R) were also assessed, however, as they were largely consistent with cFS and cFL, they are not displayed separately in the tables to avoid redundancy. A detailed version of the tables is provided in Supplementary Tables 1 and 2.

**Table 3:**
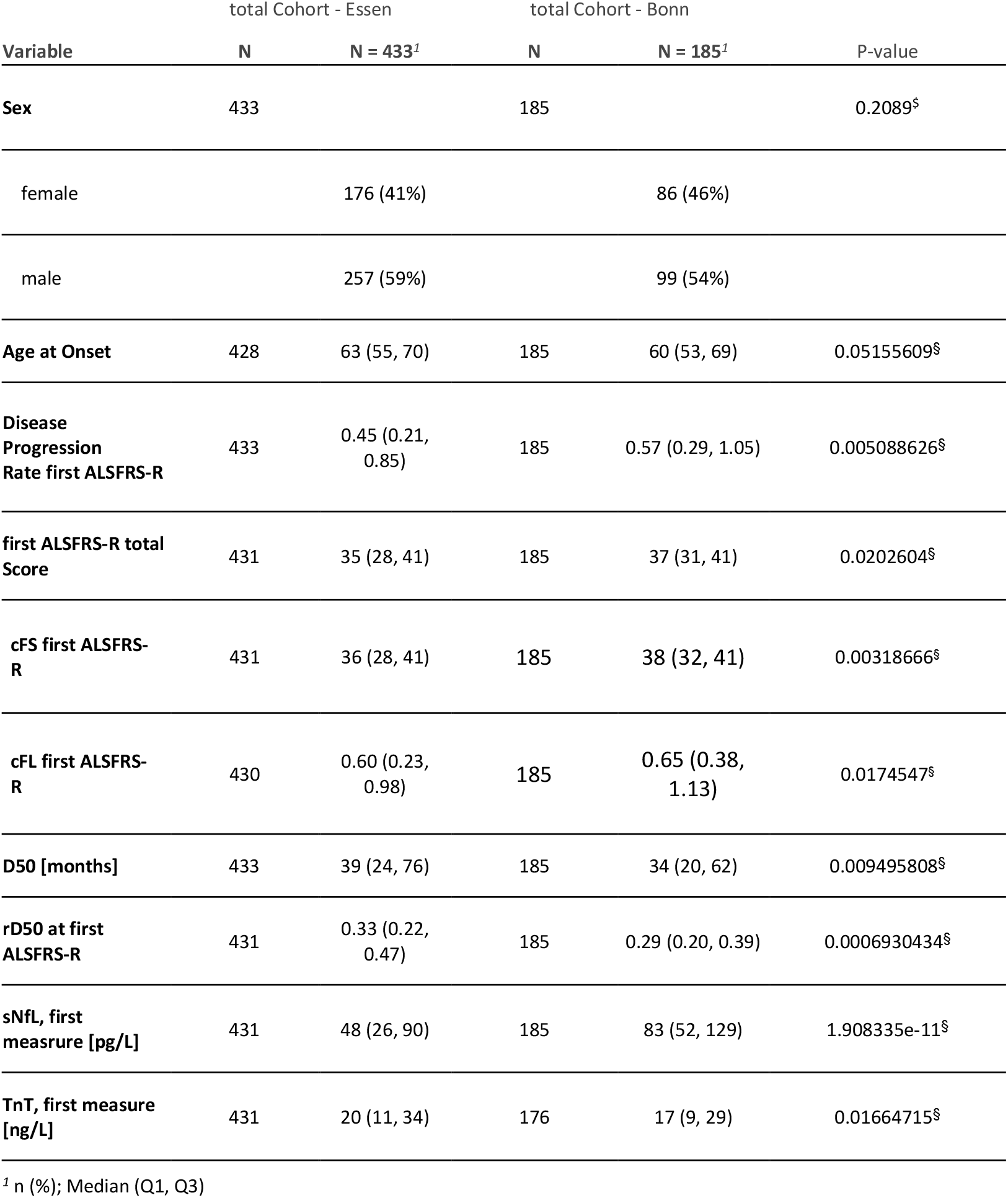
Comparison of discovery and validation cohort in relevant demographics.

| Variable | total Cohort - Essen |  | total Cohort - Bonn |  | P-value |
| --- | --- | --- | --- | --- | --- |
|  | N | N = 433<sup>1</sup> | N | N = 185<sup>1</sup> |  |
| **Sex** | 433 |  | 185 |  | 0.2089<sup>§</sup> |
| female |  | 176 (41%) |  | 86 (46%) |  |
| male |  | 257 (59%) |  | 99 (54%) |  |
| **Age at Onset** | 428 | 63 (55, 70) | 185 | 60 (53, 69) | 0.05155609<sup>§</sup> |
| **Disease Progression Rate first ALSFRS-R** | 433 | 0.45 (0.21, 0.85) | 185 | 0.57 (0.29, 1.05) | 0.005088626<sup>§</sup> |
| **first ALSFRS-R total Score** | 431 | 35 (28, 41) | 185 | 37 (31, 41) | 0.0202604<sup>§</sup> |
| **cFS first ALSFRS-R** | 431 | 36 (28, 41) | 185 | 38 (32, 41) | 0.00318666<sup>§</sup> |
| **cFL first ALSFRS-R** | 430 | 0.60 (0.23, 0.98) | 185 | 0.65 (0.38, 1.13) | 0.0174547<sup>§</sup> |
| **D50 [months]** | 433 | 39 (24, 76) | 185 | 34 (20, 62) | 0.009495808<sup>§</sup> |
| **rD50 at first ALSFRS-R** | 431 | 0.33 (0.22, 0.47) | 185 | 0.29 (0.20, 0.39) | 0.0006930434<sup>§</sup> |
| **sNfL, first measrure [pg/L]** | 431 | 48 (26, 90) | 185 | 83 (52, 129) | 1.908335e-11<sup>§</sup> |
| **TnT, first measure [ng/L]** | 431 | 20 (11, 34) | 176 | 17 (9, 29) | 0.01664715<sup>§</sup> |

<sup>1</sup> n (%); Median (Q1, Q3)

The discovery cohort showed greater functional impairment reflected by lower median ALSFRS-R scores (p=0.02) and cFS (p=0.003) while having a slower rate of disease progression with both lower DPR (p=0.005) and cFL (p=0.0017) compared to the validation cohort. The median D50 in the discovery cohort was 39 months (IQR 24–76), compared with 34 months (IQR 20–62) in the validation cohort (p = 0.009).

A higher proportion of patients was classified into the low-aggressiveness subgroup in the discovery cohort compared with the validation cohort (48% vs. 40%). In line with this, median rD50 at first sampling was higher in the discovery than in the validation cohort (0.33 [0.22–0.47] vs. 0.29 [0.20–0.39]) (p=0.0006), resulting in a different distribution of rD50-defined disease phases≐ More patients presented in advanced phases (III/IV) in the discovery cohort (21% vs. 9.3%).

ALS patients with low disease aggressiveness (high D50 value) were in an earlier phase of disease accumulation than patients with a high disease aggressiveness and low D50 value (sample shift) ^15 17^.

### 3.2 Biomarkers and disease aggressiveness (D50)

Across both cohorts, ANCOVA demonstrates a strong association of sNfL with disease aggressiveness and of TnT with disease accumulation (both p < 0.0001). Additional covariates, including age and site of onset for sNfL and sex for TnT, showed less consistent statistical significance and smaller effect sizes, generally accompanied by higher p values and lower F values (Supplementary Table 3).

#### sNfL

In the discovery cohort, ln(sNfL) concentrations at both first and last sampling time points demonstrated a strong, significant inverse correlation with D50 applied across all disease aggressiveness subgroups (p<0.001) and correlated significantly for intermediate and low disease aggressiveness subgroups in the validation cohort (Fig. 1A).

**Figure 1:**
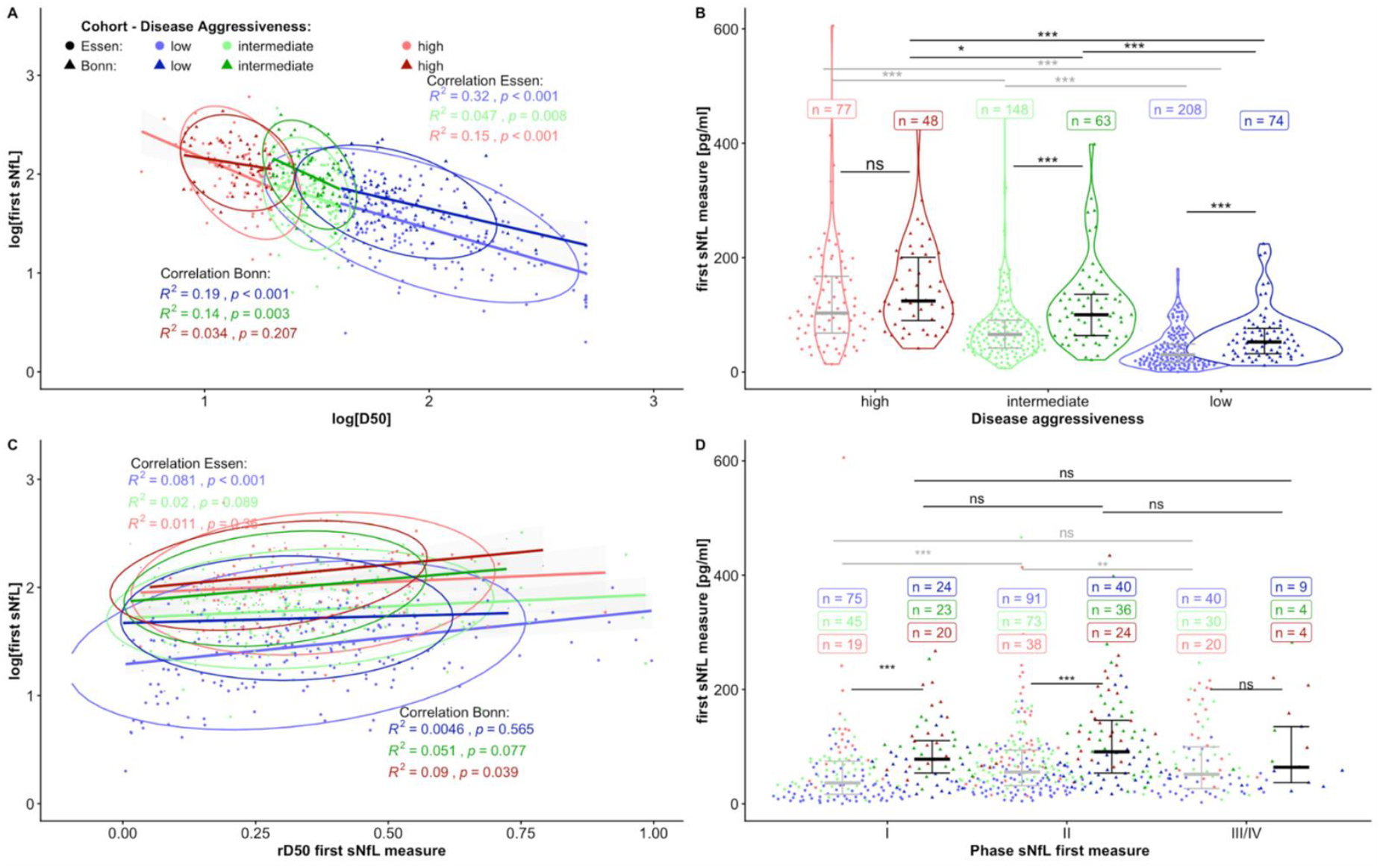
Discovery Cohort (Essen, light Colors and circles, n=433) and Validation Cohort (Bonn, dark colors and triangles, n= 185) with serum NfL (sNfL) at first measure. **(A)** Correlation of log[first_sNFL] and log[D50] with p-values and R^2^ – values as well as circles as 95% Confidence-interval and lines for linear regression for disease aggressiveness. **(B)** First measured sNFL splitted into disease aggressiveness with n as count for the individual patient in the aggressiveness subgroups and indication of significance levels within each cohort and comparing the subgroups from the two cohorts (low-low, intermediate-intermediate, high-high). **(C)** Correlation of log[first_sNfL] and rD50 at timepoint of first sNfL measure, with p- and R^2^-values, as well as circles as 95% confidence-interval and lines for linear regression for rD50 and log[first sNfL]. **(D)** First measured sNfL in rD50-derived disease phases (Phase I, Phase II, Phase III/IV with n as count of each disease aggressiveness patient in each phase). B/D Wilcoxon-rank-sum-test with Bonferroni p-value adjustment within each cohort and wilcoxon-testing with Bonferroni p-value adjustment as comparison the subgroups from each cohort and indication of significance levels within each cohort and comparing the subgroups from the two cohorts (Phase I-I, Phase II-II, Phase III/IV-III/IV). Colors for disease aggressiveness in general: high = red, D50 < 20 months; intermediate = green, D50 20-40 months; low = blue, D50 > 40 months.

The comparison of sNfL concentrations between the D50-derived disease aggressiveness subgroups revealed a statistically significant difference between all aggressiveness subgroups (p<0.005) in the discovery cohort. In the validation cohort the disease aggressiveness subgroups differed significantly, but weaker in the high vs. intermediate aggressiveness subgroup at first and last sampling (p<0.05) (Fig.1B).

#### TnT

Associations between ln(TnT) levels and D50 disease aggressiveness groups were generally weaker and less consistent between the two cohorts. In the discovery cohort only low aggressiveness subgroups correlated significantly (p<0.001), intermediate aggressiveness subgroups showed a weak correlation (p<0.05) with ln(TnT) levels. High disease aggressiveness subgroups showed no significant correlation with ln(TnT). There was no significant correlation in the validation cohort between ln(TnT) across all disease subgroups at first and last sampling except a weak correlation between ln(TnT) at last sampling with the low disease aggressiveness group (p=0.044) (Fig. 2A).

**Figure 2:**
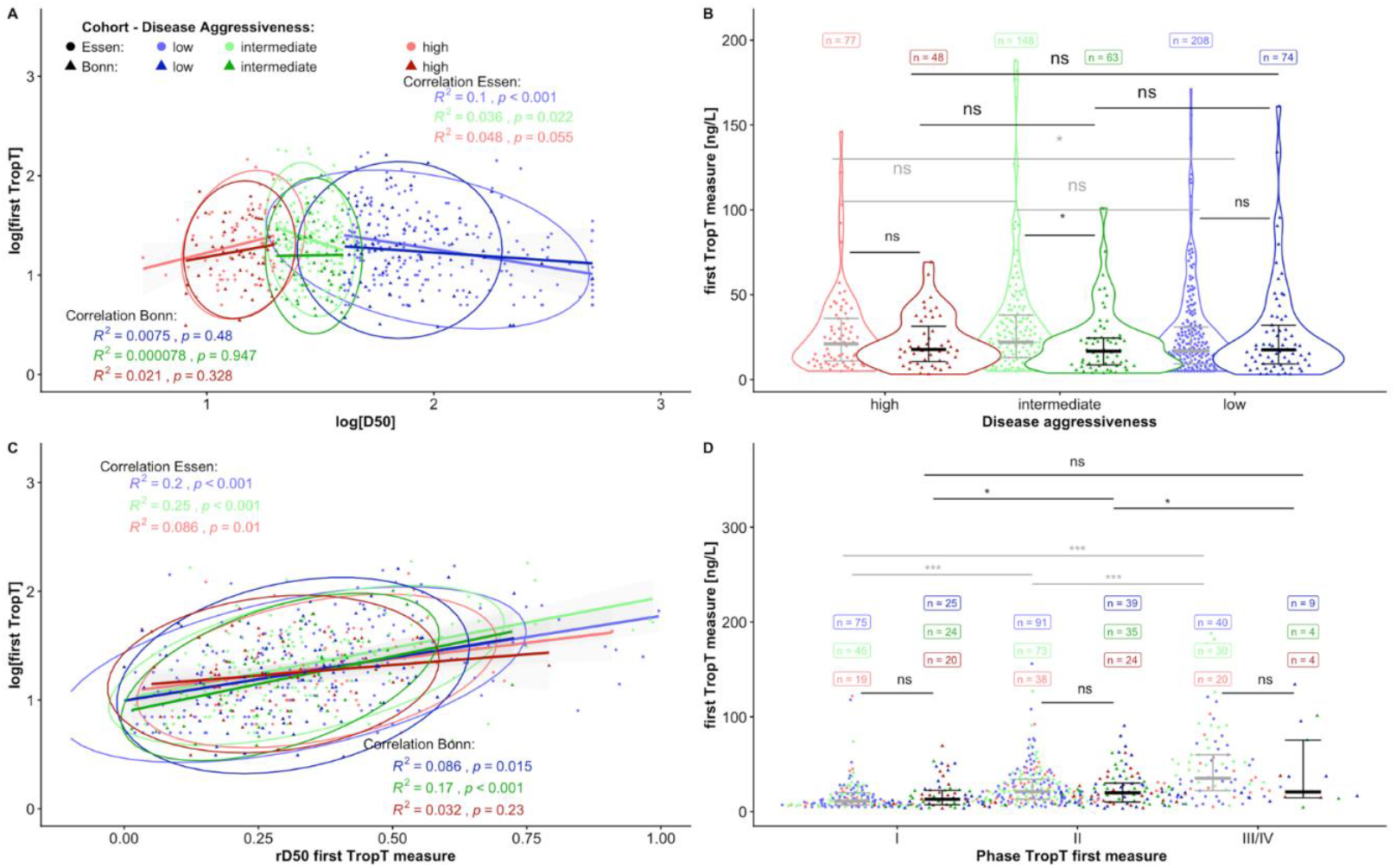
Discovery Cohort (Essen, light Colors and circles, n = 433) and Validation Cohort (Bonn, dark Colors and triangles, n = 185) Troponin T (TnT) at first measure. **(A)** Correlation of log[first_TnT] and log[D50], with p-values and R^2^-values, as well as circles as 95% Confidence-interval and Lines for Linear regression for log [D50] and log [first TnT] splitted into disease aggressiveness. **(B)** First measured TnT Levels splitted into disease aggressiveness with n as count the individual patient in the aggressiveness subgroups and indication of significance levels within each Cohort and comparing the subgroups from the two cohorts (low-low, intermediate-intermediate, high-high). **(C)** Correlation of log[first_TnT] and rD50 at timepoint of first sNfL measure, with p-values and R^2^-values, as well as circles as 95% Confidence-interval and Lines for Linear regression for rD50 and log[first TnT] splitted into disease aggressiveness. **(D)** First measured TnT in rD50-derived disease phases (Phase I, Phase II, Phase III/IV) with n as count of each disease aggressivness patient in each phase for both cohorts and indication of significance levels within each Cohort and comparing the subgroups from the two cohorts (Phase I-I, Phase II-II, Phase III/IV-III/IV). B/D Wilcoxon-rank-sum-test with Bonferroni p-value adjustment within each cohort and wilcoxon-testing with Bonferroni p-value adjustment as comparison the subgroups from each cohort. Colors for disease aggressiveness: high = red, D50 < 20 months; intermediate = green, D50 20-40 months; low = blue, D50 > 40 months.

TnT levels in the discovery cohort showed no statistically significant difference between the D50-derived disease aggressiveness subgroups, except between intermediate and low aggressiveness groups at first (p=0.02) and last (p<0.001) sampling point. In the validation cohort there was no statistically significant difference between TnT and disease aggressiveness (Fig. 2B).

### 3.3 Biomarkers and disease accumulation (rD50)

#### sNfL

There was no consistent pattern regarding sNfL and the rD50 phases. The discovery cohort showed no significant correlation of ln(sNfL) and rD50-defined disease phases with intermediate and high aggressiveness disease subgroups (p > 0.05) at first and last sample collection. In the low aggressiveness subgroup ln(sNfL) correlated significantly with rD50-phases at both sampling time points (p<0.001) (Fig. 1C). When compared between rD50-phases, sNfL demonstrated a statistically significant difference between phases I and II (p<0.001) and phases I and III/IV at both sampling points but no statistically significant difference between phases II and III/IV at first and last sampling in the discovery cohort (Fig. 1D).

In the validation cohort, ln(sNfL) showed only a weak correlation with rD50 phases within the high-aggressiveness subgroup at both first (p = 0.039) and last sampling (p = 0.027), whereas no significant associations were observed in the intermediate or low-aggressiveness subgroups (Fig. 1C).

sNfL levels differed significantly when comparing phase I with phase II and phase I with phases III/IV at first and last sampling point (p < 0.001). However, no statistically significant differences in sNfL levels were observed between disease phases II and III/IV at first and last sample collection (Fig. 1D).

#### TnT

In the discovery cohort, ln(TnT) demonstrated a robust correlation with rD50-defined phases across all aggressiveness subgroups at both sampling points (Fig. 2C). There was a statistically significant difference in-between all rD50-phases with TnT at first and last collection time points (p<0.001) (Fig. 2D).

These findings were partially replicated in the validation cohort, as there was a statistically significant correlation with intermediate and low aggressiveness subgroups at first sampling, but not in the high aggressiveness group. At last sampling the low aggressiveness group failed to reach statistical significance (p=0.06), while the intermediate subgroup remained statistically significant (p<0.001) (Fig. 2C).

There was a statistically significant difference between TnT and phases I and II (p=0.01) and I and III (p=0.01) at first sampling and between phases II and III/IV (p=0.02) at last sampling (Fig. 2D).

Of note, Fig. 1 and 2 display only data from the initial sampling tine point; data from the final sampling collection are not shown.

## 4. Discussion

We investigated the relationship between TnT, an emerging ALS biomarker, with D50-derived quantitative disease course parameters and evaluated its informative value relative to sNfL, an established marker of neurodegeneration.

Our findings demonstrate a strong and consistent association between TnT and disease accumulation across all rD50 disease phases in the discovery cohort, independent of D50-defined disease aggressiveness subgroups. Given the temporal dynamics of TnT levels, with a progressive increase over the disease course ^21 22 27 28^, an association with cumulative disease burden would be expected and is supported by our results.

This is in line with previous studies demonstrating an association between lower motor neuron and muscle derived parameters, such as the motor unit number index (MUNIX), and disease accumulation within the D50 model ^29 30^.

Although the considerably smaller validation cohort did not fully replicate all correlations observed in the discovery cohort - potentially due to limited statistical power in smaller subgroups - the overall concordance between the two cohorts strongly supports the robustness of the observed associations.

Concerning disease aggressiveness, there is only a significant correlation of TnT with the low aggressiveness group in the discovery cohort, which is weaker for the intermediate subgroup. This finding could not be replicated in the validation cohort, there was only a correlation for TnT with the low subgroup at first sampling.

These differences may reflect cohort-specific characteristics, as the low aggressiveness subgroup in the discovery cohort demonstrates a less aggressive disease course, characterized by both lower DPR and cFL as well as higher D50 values.

These findings support the interpretation that lower TnT levels may indicate a less aggressive disease course; however, TnT alone lacks sufficient discriminatory capacity to reliably differentiate among the remaining aggressiveness subgroups. In this context, Lindenborn et al.^20^ reported that patients with ALS exhibiting concurrently low TnT and sNfL levels represent a distinct subgroup with a minimally aggressive phenotype.

Overall, absolute TnT levels do not prove to be a direct predictor for disease aggressiveness, as expected.

Neurofilaments in CSF have been shown to be associated with disease aggressiveness but not disease accumulation ^10 15^. Although this relationship has not yet been explicitly examined for sNfL, it is plausible to assume that sNfL exhibits a similar pattern. Analysis of sNfL revealed an excellent correlation with disease aggressiveness (D50) across all subgroups in the discovery cohort, a finding that was confirmed in the validation cohort. These results are consistent with previous studies demonstrating a relationship between CSF-NfL and disease aggressiveness within the context of the D50 model. Our finding that this correlation extends to serum NfL levels has significant practical implications for monitoring neurofilaments in ALS.In contrast, the association between sNfL and disease accumulation, as defined by rD50-derived phases, was less clear. Meyer et al. 2025 and Dreger at al. 2021 have already shown that CSF-NfL levels do not correlate with disease burden and the rD50 disease phases. This observation is consistent with our findings for sNfL and is further supported by our data.

In the discovery cohort however, a modest relationship for sNfL was observed in the low-aggressiveness subgroup and between the early disease phase and the later phases (II and III/IV), but this pattern could not be reproduced in the validation cohort. The absence of a stable significant effect of the disease phases on sNfL levels rather indicate that sNfL concentrations remain longitudinally stable throughout the disease course, especially for the later phases II and III/IV. Whether this finding represents random variation or a true difference between CSF and serum NfL remains to be determined, highlighting the need for further studies to clarify the relationship. A possible explanation is that the discovery cohort is much larger and has substantially more patients in phase I with a low aggressive disease course than in phases II and III/IV. This suggests the presence of a cohort specific subgroup with a slower and less aggressive disease course which contributed to the apparent difference between the phases. Notably, this difference was not observed in the validation cohort.

Although sNfL concentrations were obtained using different assay platforms across cohorts (SIMOA vs ELISA), this is unlikely to have substantially influenced the results, as ELISA-related imprecision predominantly affects at low concentration ranges (<10-20 pg/mL) that are infrequently observed in ALS. Moreover, the consistency of findings across differently assessed cohorts further supports the robustness of the observed associations.

A limitation of this study is its observational design. To mitigate this, we included a validation cohort from an independent ALS center, although this cohort was smaller and not perfectly matched, yet this is taken into account by D50 stratification. Extending the analysis to include additional clinical variables, such as upper and lower motor neuron signs, phenotypes, and genetic subtypes, would be informative; however, the availability of these data was limited by the retrospective nature of the study. Prospective studies with larger, well-characterized cohorts will be essential to validate and further extend these findings.

In conclusion, our findings demonstrate that TnT levels, when interpreted within the D50 framework, constitute a robust biomarker of individual disease accumulation in patients with ALS across all disease phases, independent of disease aggressiveness. TnT thus emerges as a reliable indicator of cumulative disease burden and a promising candidate for longitudinal monitoring.

Moreover, our results support the concept that TnT reflects a distinct aspect of ALS pathology, likely linked to lower motor neuron or muscle involvement, whereas sNfL predominantly captures axonal stress. Taken together, the complementary nature of TnT and sNfL highlights the value of an integrated biomarker approach for more refined disease stratification.

Consequently, combining both biomarkers may not only guide clinical decision-making but also refine disease course characterization, monitor therapy response, and optimize the design of clinical trial cohorts, leveraging complementary insights into ALS biology.

## Data Availability

All data produced in the present study are available upon reasonable request to the authors

## List of Abbreviations

ALS: Amyotrophic lateral sclerosis
ALSFRS-R: Amyotrophic Lateral Sclerosis Functional Rating Scale-Revised
ANCOVA: Analysis of Covariance
cFL: Calculated functional loss rate
cFS: Calculated functional state
CSF: cerebrospinal fluid
TnT: Troponin T
D50: Time in months from symptom onset to 50 percent functional loss
DPR: Disease progression rate
ECLIA: Electrochemiluminescence Immunoassay
ELISA: Enzyme-linked immunosorbent assay
IQR: Interquartile range
In: Natural logarithm
MUNIX: Motor unit number index
NfL: Neurofilament light chain
CSF-NfL: Neurofilament light chain (measured in cerebrospinal fluid)
sNfL: Neurofilament light chain (measured in serum)
pNfH: Phosphorylated neurofilament heavy chain
rD50: Relative D50
SIMOA: Single molecule array

## REFERENCES

1 Feldman EL, Goutman SA, Petri S, et al. Amyotrophic lateral sclerosis. Lancet 2022;400:1363–80. doi: 10.1016/S0140-6736(22)01272-7

2 Chiò A, Calvo A, Moglia C, Mazzini L, Mora G. Phenotypic heterogeneity of amyotrophic lateral sclerosis: a population based study. J Neurol Neurosurg Psychiatry 2011;82:740–46. doi: 10.1136/jnnp.2010.235952

3 Chia R, Moaddel R, Kwan JY, et al. A plasma proteomics-based candidate biomarker panel predictive of amyotrophic lateral sclerosis. Nat Med 2025;31:3440–50. doi: 10.1038/s41591-025-03890-6

4 Cedarbaum JM, Stambler N, Malta E, et al. The ALSFRS-R: a revised ALS functional rating scale that incorporates assessments of respiratory function. BDNF ALS Study Group (Phase III). J Neurol Sci 1999;169:13–21. doi: 10.1016/s0022-510x(99)00210-5

5 Paganoni S, Cudkowicz M, Berry JD. Outcome measures in amyotrophic lateral sclerosis clinical trials. Clin Investig (Lond) 2014;4:605–18. doi: 10.4155/cli.14.52

6 van Eijk RPA, Jongh AD de, Nikolakopoulos S, et al. An old friend who has overstayed their welcome: the ALSFRS-R total score as primary endpoint for ALS clinical trials. Amyotroph Lateral Scler Frontotemporal Degener 2021;22:300–07. doi: 10.1080/21678421.2021.1879865

7 van Eijk RPA, Weemering DN, Opie-Martin S, et al. Natural history of the revised ALS functional rating scale and its association with survival: the PRECISION-ALS Extant Study. Amyotroph Lateral Scler Frontotemporal Degener 2025;26:30–40. doi: 10.1080/21678421.2024.2443985

8 Bakker LA, Schröder CD, Tan HHG, et al. Development and assessment of the inter-rater and intra-rater reproducibility of a self-administration version of the ALSFRS-R. J Neurol Neurosurg Psychiatry 2020;91:75–81. doi: 10.1136/jnnp-2019-321138

9 Proudfoot M, Jones A, Talbot K, Al-Chalabi A, Turner MR. The ALSFRS as an outcome measure in therapeutic trials and its relationship to symptom onset. Amyotroph Lateral Scler Frontotemporal Degener 2016;17:414–25. doi: 10.3109/21678421.2016.1140786

10 Meyer J, Gaur N, Gablentz J von der, et al. Phosphorylated neurofilament heavy chain (pNfH) concentration in cerebrospinal fluid predicts overall disease aggressiveness (D50) in amyotrophic lateral sclerosis. Front Neurosci 2025;19:1536818. doi: 10.3389/fnins.2025.1536818

11 Poesen K, Schaepdryver M de, Stubendorff B, et al. Neurofilament markers for ALS correlate with extent of upper and lower motor neuron disease. Neurology 2017;88:2302– 09. doi: 10.1212/WNL.0000000000004029

12 Benatar M, Macklin EA, Malaspina A, et al. Prognostic clinical and biological markers for amyotrophic lateral sclerosis disease progression: validation and implications for clinical trial design and analysis. EBioMedicine 2024;108:105323. doi: 10.1016/j.ebiom.2024.105323

13 Poesen K, van Damme P. Diagnostic and Prognostic Performance of Neurofilaments in ALS. Front Neurol 2018;9:1167. doi: 10.3389/fneur.2018.01167

14 Gaetani L, Blennow K, Calabresi P, Di Filippo M, Parnetti L, Zetterberg H. Neurofilament light chain as a biomarker in neurological disorders. J Neurol Neurosurg Psychiatry 2019;90:870–81. doi: 10.1136/jnnp-2018-320106

15 Dreger M, Steinbach R, Gaur N, et al. Cerebrospinal Fluid Neurofilament Light Chain (NfL) Predicts Disease Aggressiveness in Amyotrophic Lateral Sclerosis: An Application of the D50 Disease Progression Model. Front Neurosci 2021;15:651651. doi: 10.3389/fnins.2021.651651

16 Steinbach R, Gaur N, Roediger A, et al. Disease aggressiveness signatures of amyotrophic lateral sclerosis in white matter tracts revealed by the D50 disease progression model. Hum Brain Mapp 2021;42:737–52. doi: 10.1002/hbm.25258

17 Dreger M, Steinbach R, Otto M, Turner MR, Grosskreutz J. Cerebrospinal fluid biomarkers of disease activity and progression in amyotrophic lateral sclerosis. J Neurol Neurosurg Psychiatry 2022;93:422–35. doi: 10.1136/jnnp-2021-327503

18 Vidovic M, Lapp HS, Weber C, et al. Comparative analysis of neurofilaments and biomarkers of muscular damage in amyotrophic lateral sclerosis. Brain Commun 2024;6:fcae288. doi: 10.1093/braincomms/fcae288

19 Castro-Gomez S, Radermacher B, Tacik P, Mirandola SR, Heneka MT, Weydt P. Teaching an old dog new tricks: serum troponin T as a biomarker in amyotrophic lateral sclerosis. Brain Commun 2021;3:fcab274. doi: 10.1093/braincomms/fcab274

20 Lindenborn P, Fabian R, Grehl T, et al. Combination of serum neurofilament light chain and serum cardiac troponin T as biomarkers improves diagnostic accuracy in amyotrophic lateral sclerosis, 2025.

21 Koch T, Fabian R, Weinhold L, et al. Cardiac troponin T as a serum biomarker of respiratory impairment in amyotrophic lateral sclerosis. Ann Clin Transl Neurol 2024;11:2063–72. doi: 10.1002/acn3.52126

22 Bernsen S, Fabian R, Koc Y, et al. Serum Cardiac Troponin T Levels as a Therapy Response Marker in Tofersen-Treated ALS. Muscle Nerve 2025. doi: 10.1002/mus.28453

23 Steinacker P, Feneberg E, Weishaupt J, et al. Neurofilaments in the diagnosis of motoneuron diseases: a prospective study on 455 patients. J Neurol Neurosurg Psychiatry 2016;87:12–20. doi: 10.1136/jnnp-2015-311387

24 Gaur N, Steinbach R, Plaas M, Witte OW, Brill MS, Grosskreutz J. Chitinase dysregulation predicts disease aggressiveness in ALS: Insights from the D50 progression model. J Neurol Neurosurg Psychiatry 2023;94:585–88. doi: 10.1136/jnnp-2022-330318

25 Steinbach R, Batyrbekova M, Gaur N, et al. Applying the D50 disease progression model to gray and white matter pathology in amyotrophic lateral sclerosis. Neuroimage Clin 2020;25:102094. doi: 10.1016/j.nicl.2019.102094

26 Prell T, Steinbach R, Witte OW, Grosskreutz J. Poor emotional well-being is associated with rapid progression in amyotrophic lateral sclerosis. eNeurologicalSci 2019;16:100198. doi: 10.1016/j.ensci.2019.100198

27 Kläppe U, Chamoun S, Shen Q, et al. Cardiac troponin T is elevated and increases longitudinally in ALS patients. Amyotroph Lateral Scler Frontotemporal Degener 2022;23:58– 65. doi: 10.1080/21678421.2021.1939384

28 Chamoun S, Imrell S, Upate Z, et al. Plasma troponin T reflects lower motor neuron involvement on electromyography in amyotrophic lateral sclerosis. Brain Commun 2025;7:fcaf177. doi: 10.1093/braincomms/fcaf177

29 Ebersbach T, Roediger A, Steinbach R, et al. Motor unit number index (MUNIX) loss of 50% occurs in half the time of 50% functional loss according to the D50 disease progression model of ALS. Sci Rep 2023;13:3981. doi: 10.1038/s41598-023-30871-x

30 Ebersbach T, Roediger A, Steinbach R, et al. Motor unit number index (MUNIX) in the D50 disease progression model reflects disease accumulation independently of disease aggressiveness in ALS. Sci Rep 2022;12:15997. doi: 10.1038/s41598-022-19911-0

