## Supplementary material for "Troponin T and Neurofilament Light Chain Levels as Complementary Biomarkers of Disease Accumulation and Aggressiveness in Amyotrophic Lateral Sclerosis": -

**Supplementary Table 1:** Demographic and clinical characteristics of the discovery cohort, full version (Essen, n = 433)

| **Variable** | total Cohort - Essen | | D50 Disease Aggressiveness | | | |
| --- | --- | --- | --- | --- | --- | --- |
|  | **N** | **N = 433***^1^* | **high** N = 77*^1^* | **intermediate** N = 148*^1^* | **low** N = 208*^1^* | **p-value***^2^* |
| **Sex** | 433 |  |  |  |  | 0.11 |
| female |  | 176 (41%) | 38 (49%) | 63 (43%) | 75 (36%) |  |
| **Age at Onset** | 428 | 63 (55, 70) | 64 (57, 71) | 66 (56, 73) | 60 (54, 68) | <0.001 |
| **Symptom onset Region** | 433 |  |  |  |  | 0.002 |
| bulbar |  | 70 (16%) | 14 (18%) | 34 (23%) | 22 (11%) |  |
| spinal |  | 329 (76%) | 53 (69%) | 101 (68%) | 175 (84%) |  |
| unknown |  | 34 (7.9%) | 10 (13%) | 13 (8.8%) | 11 (5.3%) |  |
| **first ALSFRS-R Time since onset [months]** | 431 | 25 (13, 50) | 10 (7, 14) | 19 (13, 29) | 47 (29, 97) | <0.001 |
| **last ALSFRSR Time since onset [months]** | 431 | 31 (18, 57) | 13 (8, 16) | 25 (19, 34) | 55 (37, 99) | <0.001 |
| **first ALSFRS-R total score** | 431 | 35 (28, 41) | 32 (27, 38) | 35 (27, 40) | 36 (29, 43) | 0.004 |
| **last ALSFRS-R total score** | 431 | 32 (24, 39) | 28 (22, 33) | 30 (23, 37) | 35 (27, 42) | <0.001 |
| **Disease Progression Rate first ALSFRS-R** | 433 | 0.45 (0.21, 0.85) | 1.43 (1.10, 2.00) | 0.63 (0.47, 0.82) | 0.21 (0.12, 0.36) | <0.001 |
| **Disease Progression Rate last ALSFRS-R** | 427 | 0.51 (0.22, 0.90) | 1.38 (1.18, 1.88) | 0.73 (0.59, 0.87) | 0.20 (0.09, 0.34) | <0.001 |
| **cFS first ALSFRS-R** | 431 | 36 (28, 41) | 33 (29, 39) | 36 (28, 41) | 36 (28, 43) | 0.052 |
| **cFL first ALSFRS-R** | 430 | 0.60 (0.23, 0.98) | 1.61 (1.33, 1.97) | 0.82 (0.66, 0.96) | 0.23 (0.12, 0.43) | <0.001 |
| **cFS last ALSFRS-R** | 431 | 31 (23, 38) | 26 (22, 35) | 30 (22, 35) | 34 (26, 41) | <0.001 |
| **cFL last ALSFRS-R** | 430 | 0.65 (0.25, 1.14) | 1.63 (1.36, 2.15) | 0.91 (0.73, 1.14) | 0.25 (0.12, 0.45) | <0.001 |
| **D50 [months]** | 433 | 39 (24, 76) | 15 (12, 18) | 28 (24, 34) | 81 (54, 174) | <0.001 |
| **rD50 at first ALSFRS-R** | 431 | 0.33 (0.22, 0.47) | 0.37 (0.25, 0.44) | 0.33 (0.23, 0.47) | 0.32 (0.18, 0.47) | 0.2 |
| **disease phase first ALSFRS-R** | 431 |  |  |  |  | 0.4 |
| I |  | 139 (32%) | 19 (25%) | 45 (30%) | 75 (36%) |  |
| II |  | 203 (47%) | 40 (52%) | 72 (49%) | 91 (44%) |  |
| III/IV |  | 89 (21%) | 18 (23%) | 31 (21%) | 40 (19%) |  |
| **rD50 at last ALSFRS-R** | 431 | 0.42 (0.29, 0.54) | 0.50 (0.35, 0.56) | 0.44 (0.34, 0.56) | 0.37 (0.22, 0.51) | <0.001 |
| **disease phase last ALSFRS-R** | 431 |  |  |  |  | <0.001 |
| I |  | 91 (21%) | 11 (14%) | 18 (12%) | 62 (30%) |  |
| II |  | 193 (45%) | 28 (36%) | 76 (51%) | 89 (43%) |  |
| III/IV |  | 147 (34%) | 38 (49%) | 54 (36%) | 55 (27%) |  |
| **sNfL, first measure [pg/ml]** | 431 | 49 (27, 90) | 103 (68, 167) | 66 (42, 91) | 30 (17, 49) | <0.001 |
| **rD50 first sNfL measure** | 431 | 0.33 (0.22, 0.47) | 0.37 (0.25, 0.50) | 0.33 (0.23, 0.47) | 0.32 (0.17, 0.47) | 0.2 |
| **cFL first sNfL measure** | 430 | 0.60 (0.24, 0.98) | 1.61 (1.31, 1.97) | 0.82 (0.66, 0.97) | 0.24 (0.12, 0.43) | <0.001 |
| **cFS first sNfL measure** | 431 | 36 (28, 41) | 33 (26, 39) | 36 (28, 41) | 36 (28, 43) | 0.059 |
| **sNfL, last measure [pg/ml]** | 431 | 53 (28, 96) | 111 (74, 179) | 73 (44, 102) | 31 (16, 51) | <0.001 |
| **rD50 last sNfL measure** | 431 | 0.41 (0.28, 0.54) | 0.43 (0.30, 0.56) | 0.44 (0.33, 0.55) | 0.37 (0.22, 0.50) | 0.001 |
| **cFL last sNfL measure** | 430 | 0.64 (0.25, 1.14) | 1.63 (1.34, 2.15) | 0.91 (0.72, 1.14) | 0.25 (0.12, 0.47) | <0.001 |
| **cFS last sNfL measure** | 431 | 32 (24, 38) | 30 (23, 36) | 30 (23, 36) | 34 (26, 41) | <0.001 |
| **TnT, first measure [ng/L]** | 431 | 20 (11, 34) | 21 (11, 36) | 22 (13, 38) | 17 (9, 31) | 0.023 |
| **rD50 first TnT measure** | 431 | 0.33 (0.22, 0.47) | 0.37 (0.25, 0.50) | 0.33 (0.23, 0.47) | 0.32 (0.18, 0.47) | 0.2 |
| **cFL first TnT measure** | 430 | 0.60 (0.23, 0.98) | 1.61 (1.33, 1.97) | 0.82 (0.66, 0.96) | 0.23 (0.12, 0.43) | <0.001 |
| **cFS first TnT measure** | 431 | 36 (28, 41) | 33 (26, 39) | 36 (28, 41) | 36 (28, 43) | 0.046 |
| **TnT, last measure [ng/L]** | 431 | 23 (12, 39) | 22 (12, 39) | 29 (18, 43) | 20 (10, 34) | <0.001 |
| **rD50 last TnT measure** | 431 | 0.41 (0.28, 0.54) | 0.43 (0.31, 0.56) | 0.44 (0.33, 0.55) | 0.37 (0.22, 0.50) | <0.001 |
| **cFL last TnT measure** | 430 | 0.65 (0.25, 1.14) | 1.63 (1.35, 2.15) | 0.91 (0.73, 1.14) | 0.25 (0.12, 0.45) | <0.001 |
| **cFS last TnT measure** | 431 | 31 (23, 38) | 30 (23, 36) | 30 (23, 36) | 34 (26, 41) | <0.001 |
| *^1^* n (%); Median (Q1, Q3) | | | | | | |
| *^2^* Pearson’s Chi-squared test; Kruskal-Wallis rank sum test | | | | | | |
| ALSFRS-R = Amyotrophic Lateral Sclerosis Rating Scale-Revised; cFL= calculated Functional Loss rate; cFS= calculated Functional State, n/N = Number; sNfL = serum Neurofilament light chain; TnT = Troponin T | | | | | | |
| rD50, cFL and cFS first and last measure for ALSFRS-R, sNFL and TnT | | | | | | |

**Supplementary Table 2:** Demographic and clinical characteristics of the validation cohort, full version (Bonn)

| **Variable** | total Cohort - Bonn | | splitted into D50 Disease Aggressiveness | | | |
| --- | --- | --- | --- | --- | --- | --- |
|  | **N** | **N = 185***^1^* | **high** N = 48*^1^* | **intermediate** N = 63*^1^* | **low** N = 74*^1^* | **p-value***^2^* |
| **Sex** | 185 |  |  |  |  | >0.9 |
| female |  | 86 (46%) | 22 (46%) | 30 (48%) | 34 (46%) |  |
| **Age at Onset, years** | 185 | 60 (53, 69) | 64 (55, 73) | 62 (56, 70) | 58 (51, 66) | 0.018 |
| **Symptom Onset Region** | 185 |  |  |  |  | 0.050 |
| bulbar |  | 42 (23%) | 15 (31%) | 18 (29%) | 9 (12%) |  |
| spinal |  | 133 (72%) | 31 (65%) | 43 (68%) | 59 (80%) |  |
| n.a. |  | 10 (5.4%) | 2 (4.2%) | 2 (3.2%) | 6 (8.1%) |  |
| **first ALSFRSR Time since onset [months]** | 185 | 18 (10, 35) | 7 (5, 11) | 17 (11, 20) | 45 (25, 68) | <0.001 |
| **last ALSFRSR Time since onset [months]** | 184 | 30 (19, 52) | 14 (11, 19) | 27 (21, 34) | 58 (44, 85) | <0.001 |
| **first ALSFRS-R total Score** | 185 | 37 (31, 41) | 36 (29, 41) | 38 (31, 40) | 37 (31, 41) | 0.5 |
| **last ALSFRS-R total Score** | 185 | 28 (22, 34) | 26 (19, 31) | 27 (23, 33) | 30 (25, 36) | 0.003 |
| **Disease Progression Rate first ALSFRS-R** | 185 | 0.57 (0.29, 1.05) | 1.62 (1.11, 2.54) | 0.67 (0.52, 0.92) | 0.28 (0.18, 0.39) | <0.001 |
| **Disease Progression Rate last ALSFRS-R** | 184 | 0.62 (0.33, 1.15) | 1.50 (1.26, 1.83) | 0.72 (0.59, 0.86) | 0.29 (0.18, 0.38) | <0.001 |
| **D50 [months]** | 185 | 34 (20, 62) | 14 (12, 17) | 30 (25, 35) | 64 (54, 104) | <0.001 |
| **rD50 at first ALSFRS-R** | 185 | 0.29 (0.20, 0.39) | 0.26 (0.18, 0.36) | 0.27 (0.20, 0.38) | 0.32 (0.21, 0.40) | 0.4 |
| **rD50 at last ALSFRS-R** | 184 | 0.47 (0.37, 0.57) | 0.54 (0.42, 0.65) | 0.49 (0.36, 0.56) | 0.44 (0.35, 0.52) | 0.002 |
| **Disease phase first ALSFRS-R** | 183 |  |  |  |  | 0.7 |
| I |  | 70 (38%) | 20 (43%) | 25 (40%) | 25 (34%) |  |
| II |  | 96 (52%) | 23 (49%) | 34 (54%) | 39 (53%) |  |
| III/IV |  | 17 (9.3%) | 4 (8.5%) | 4 (6.3%) | 9 (12%) |  |
| **Disease phase last ALSFRS-R** | 184 |  |  |  |  | <0.001 |
| I |  | 9 (4.9%) | 0 (0%) | 0 (0%) | 9 (12%) |  |
| II |  | 101 (55%) | 21 (45%) | 36 (57%) | 44 (59%) |  |
| III/IV |  | 74 (40%) | 26 (55%) | 27 (43%) | 21 (28%) |  |
| **sNfL, first measure [pg/ml]** | 185 | 83 (52, 129) | 124 (88, 202) | 100 (63, 137) | 53 (32, 77) | <0.001 |
| **rD50 first sNfL measure** | 185 | 0.29 (0.20, 0.39) | 0.27 (0.18, 0.36) | 0.27 (0.20, 0.38) | 0.32 (0.21, 0.40) | 0.5 |
| **cFL_sNfL_first measure** | 185 | 0.65 (0.38, 1.12) | 1.55 (1.28, 1.91) | 0.80 (0.61, 1.03) | 0.34 (0.24, 0.48) | <0.001 |
| **cFS_sNfL_first measure** | 185 | 38 (32, 41) | 38 (33, 41) | 38 (33, 42) | 37 (32, 42) | 0.9 |
| **NfL, last measure [pg/ml]** | 185 | 90 (51, 142) | 138 (98, 197) | 108 (59, 145) | 52 (30, 76) | <0.001 |
| **rD50 last sNfL measure** | 184 | 0.47 (0.38, 0.57) | 0.55 (0.43, 0.67) | 0.49 (0.37, 0.56) | 0.44 (0.35, 0.52) | <0.001 |
| **cFL_sNfL_last measure** | 184 | 0.79 (0.46, 1.35) | 1.80 (1.49, 2.16) | 0.96 (0.78, 1.14) | 0.40 (0.26, 0.51) | <0.001 |
| **cFS_sNfL_last measure** | 184 | 28 (22, 33) | 23 (17, 30) | 27 (22, 33) | 30 (25, 35) | <0.001 |
| **TnT, first measure [ng/L]** | 176 | 17 (9, 29) | 18 (10, 34) | 17 (8, 25) | 18 (9, 32) | 0.8 |
| **rD50 first TnT measure** | 185 | 0.29 (0.20, 0.39) | 0.27 (0.18, 0.36) | 0.29 (0.20, 0.38) | 0.32 (0.21, 0.40) | 0.5 |
| **cFL_TnT_first measure** | 185 | 0.65 (0.38, 1.12) | 1.55 (1.27, 1.91) | 0.80 (0.61, 1.04) | 0.34 (0.23, 0.48) | <0.001 |
| **cFS_TnT_first measure** | 185 | 38 (32, 41) | 38 (33, 41) | 39 (33, 42) | 37 (32, 42) | 0.9 |
| **TnT, last measure [ng/L]** | 179 | 23 (13, 39) | 26 (18, 39) | 26 (12, 38) | 21 (12, 43) | 0.4 |
| **rD50 last TnT measure** | 181 | 0.47 (0.38, 0.57) | 0.55 (0.43, 0.67) | 0.48 (0.37, 0.56) | 0.44 (0.36, 0.52) | <0.001 |
| **cFL_TnT_last measure** | 181 | 0.80 (0.46, 1.34) | 1.79 (1.49, 2.16) | 0.96 (0.79, 1.14) | 0.40 (0.26, 0.51) | <0.001 |
| **cFS_TnT_last measure** | 181 | 0.80 (0.46, 1.34) | 1.79 (1.49, 2.16) | 0.96 (0.79, 1.14) | 0.40 (0.26, 0.51) | <0.001 |
| *^1^* n (%); Median (Q1, Q3) | | | | | | |
| *^2^* Pearson’s Chi-squared test; Kruskal-Wallis rank sum test; Fisher’s exact test | | | | | | |
| ALSFRS-R = Amyotrophic Lateral Sclerosis Rating Scale-Revised; cFL= calculated Functional Loss rate; cFS= calculated Functional State, n/N = Number; sNfL = serum Neurofilament light chain; TnT = Troponin T | | | | | | |
| rD50, cFL and cFS first and last measure for ALSFRS-R, sNFL and TnT | | | | | | |

**Supplementary Table 3:** ANCOVA with cohorts first/last sNFL and first/last TnT with Cofactors: Age at Onset, rD50, D50, sex and Symptom Onset Region with effectsize (Eta square and omega)

| **Essen – sNfL first measure** | **DF** | **F-value** | **p-value** | **Eta sq (η^2^) [95% CI]** | **Omega sq (ϖ^2^) [95% CI]** |
| --- | --- | --- | --- | --- | --- |
| **Disease aggressiveness** | 2 | 85.1775 | < 2.2e-16 *** | 0.29 \| [0.23, 1.00] | 0.28 \| [0.22, 1.00] |
| **Sex** | 1 | 8.3313 | 0.004099 ** | 0.02 \| [0.00, 1.00] | 0.02 \| [0.00, 1.00] |
| **rD50 at first sNfL measure** | 1 | 0.0043 | 0.947633 | 1.03e-05 \| [0.00, 1.00] | 0.00 \| [0.00, 1.00] |
| **Age at Onset** | 1 | 22.7905 | 2.503e-06 *** | 0.05 \| [0.02, 1.00] | 0.05 \| [0.02, 1.00] |
| **Onset Region** | 2 | 19.8525 | 5.800e-09 *** | 0.09 \| [0.05, 1.00] | 0.08 \| [0.04, 1.00] |
| **Bonn -sNfL first measure** | **DF** | **F-value** | **p-value** | **Eta sq (η^2^) [95% CI]** | **Omega sq (ϖ^2^) [95% CI]** |
| **Disease aggressiveness** | 2 | 31.4837 | 2.005e-12 *** | 0.26 \| [0.17, 1.00] | 0.25 \| [0.16, 1.00] |
| **Sex** | 1 | 3.9650 | 0.047995 * | 0.02 \| [0.00, 1.00] | 0.02 \| [0.00, 1.00] |
| **rD50 at first sNfL measure** | 1 | 9.3601 | 0.002563 ** | 0.05 \| [0.01, 1.00] | 0.04 \| [0.01, 1.00] |
| **Age at Onset** | 1 | 7.8399 | 0.005678 ** | 0.04 \| [0.01, 1.00] | 0.04 \| [0.00, 1.00] |
| **Onset Region** | 2 | 4.7876 | 0.009442 ** | 0.05 \| [0.01, 1.00] | 0.04 \| [0.00, 1.00] |
| **Essen – TnT first measure** | **DF** | **F-value** | **p-value** | **Eta sq (η^2^) [95% CI]** | **Omega sq (ϖ^2^) [95% CI]** |
| **Disease aggressiveness** | 2 | 2.3562 | 0.09603 | 0.01 \| [0.00, 1.00] | 6.33e-03 \| [0.00, 1.00] |
| **Sex** | 1 | 33.4355 | 1.444e-08 *** | 0.07 \| [0.04, 1.00] | 0.07 \| [0.04, 1.00] |
| **rD50 at first sNfL measure** | 1 | 55.9682 | 4.384e-13 *** | 0.12 \| [0.07, 1.00] | 0.11 \| [0.07, 1.00] |
| **Age at Onset** | 1 | 6.0831 | 0.01405 * | 0.01 \| [0.00, 1.00] | 0.01 \| [0.00, 1.00] |
| **Onset Region** | 2 | 1.8960 | 0.15146 | 8.99e-03 \| [0.00, 1.00] | 4.19e-03 \| [0.00, 1.00]. |
| **Bonn – TnT first measure** | **DF** | **F-value** | **p-value** | **Eta sq (η^2^) [95% CI]** | **Omega sq (ϖ^2^) [95% CI]** |
| **Disease aggressiveness** | 2 | 1.5725 | 0.21056 | 0.02 \| [0.00, 1.00] | 6.46e-03 \| [0.00, 1.00] |
| **Sex** | 1 | 28.6319 | 2.833e-07 *** | 0.15 \| [0.07, 1.00] | 0.14 \| [0.07, 1.00] |
| **rD50 at first sNfL measure** | 1 | 20.2298 | 1.277e-05 *** | 0.11 \| [0.04, 1.00] | 0.10 \| [0.04, 1.00] |
| **Age at Onset** | 1 | 0.3544 | 0.55241 | 2.11e-03 \| [0.00, 1.00] | 0.00 \| [0.00, 1.00] |
| **Onset Region** | 2 | 3.4794 | 0.03306 * | 0.04 \| [0.00, 1.00] | 0.03 \| [0.00, 1.00] |
| **Essen – sNfL last measure** | **DF** | **F-value** | **p-value** | **Eta sq (η^2^) [95% CI]** | **Omega sq (ϖ^2^) [95% CI]** |
| **Disease aggressiveness** | 2 | 91.1007 | < 2.2e-16 *** | 0.30 \| [0.24, 1.00] | 0.30 \| [0.24, 1.00] |
| **Sex** | 1 | 6.6235 | 0.01041 * | 0.02 \| [0.00, 1.00] | 0.01 \| [0.00, 1.00] |
| **rD50 at last sNfL measure** | 1 | 1.0083 | 0.31589 | 2.41e-03 \| [0.00, 1.00] | 1.95e-05 \| [0.00, 1.00] |
| **Age at Onset** | 1 | 25.9428 | 5.333e-07 *** | 0.06 \| [0.03, 1.00] | 0.06 \| [0.03, 1.00] |
| **Onset Region** | 2 | 21.0412 | 1.964e-09 *** | 0.09 \| [0.05, 1.00] | 0.09 \| [0.05, 1.00] |
| **Bonn – sNfL last measure** | **DF** | **F-value** | **p-value** | **Eta sq (η^2^) [95% CI]** | **Omega sq (ϖ^2^) [95% CI]** |
| **Disease aggressiveness** | 2 | 24.9578 | 2.87e-10 *** | 0.22 \| [0.13, 1.00] | 0.21 \| [0.12, 1.00] |
| **Sex** | 1 | 3.0439 | 0.0827843 | 0.02 \| [0.00, 1.00] | 0.01 \| [0.00, 1.00] |
| **rD50 at last sNfL measure** | 1 | 5.1352 | 0.0246633 * | 0.03 \| [0.00, 1.00] | 0.02 \| [0.00, 1.00] |
| **Age at Onset** | 1 | 7.9714 | 0.0053001 ** | 0.04 \| [0.01, 1.00] | 0.04 \| [0.00, 1.00] |
| **Onset Region** | 2 | 7.4810 | 0.0007617 *** | 0.08 \| [0.02, 1.00] | 0.07 \| [0.01, 1.00] |
| **Essen – TnT last measure** | **DF** | **F-value** | **p-value** | **Eta sq (η^2^) [95% CI]** | **Omega sq (ϖ^2^) [95% CI]** |
| **Disease aggressiveness** | 2 | 4.2004 | 0.01563 * | 0.02 \| [0.00, 1.00] | 0.01 \| [0.00, 1.00] |
| **Sex** | 1 | 30.9361 | 4.767e-08 *** | 0.07 \| [0.03, 1.00] | 0.07 \| [0.03, 1.00] |
| **rD50 at last sNfL measure** | 1 | 47.8318 | 1.749e-11 *** | 0.10 \| [0.06, 1.00] | 0.10 \| [0.06, 1.00] |
| **Age at Onset** | 1 | 6.6201 | 0.01043 * | 0.02 \| [0.00, 1.00] | 0.01 \| [0.00, 1.00] |
| **Onset Region** | 2 | 2.3920 | 0.09270 | 0.01 \| [0.00, 1.00] | 6.49e-03 \| [0.00, 1.00] |
| **Bonn – TnT last measure** | **DF** | **F-value** | **p-value** | **Eta sq (η^2^) [95% CI]** | **Omega sq (ϖ^2^) [95% CI]** |
| **Disease aggressiveness** | 2 | 0.0062 | 0.99383 | 7.41e-05 \| [0.00, 1.00] | 0.00 \| [0.00, 1.00] |
| **Sex** | 1 | 22.9343 | 3.676e-06 *** | 0.12 \| [0.05, 1.00] | 0.11 \| [0.05, 1.00] |
| **rD50 at last sNfL measure** | 1 | 5.4919 | 0.02028 * | 0.03 \| [0.00, 1.00] | 0.03 \| [0.00, 1.00] |
| **Age at Onset** | 1 | 0.5607 | 0.45504 | 3.35e-03 \| [0.00, 1.00] | 0.00 \| [0.00, 1.00] |
| **Onset Region** | 2 | 1.8743 | 0.15668 | 0.02 \| [0.00, 1.00] | 9.89e-03 \| [0.00, 1.00] |

**Eta square** **(η^2^):** Measures the proportion of total variance in the dependent variable accounted for by an effect.

**Omega2 square** **(ϖ^2^):** A less biased estimate than (eta ^{2}), preferred for small sample sizes because it estimates the effect in the population rather than just the sample.

**Interpretation**:

**Small Effect:** (0.01 $\leq$ ES < 0.06) (Variance explained: (1 % - 6 % ))

**Medium Effect:** (0.06 $\leq$ ES < 0.14) (Variance explained: (6 % - 14 %))

**Large Effect:** (ES $\geq$ 0.14) (Variance explained: (>14 % ))
